# Extreme Weather and Healthcare Access in Malawi: The Impact of Precipitation on ANC Service Utilisation

**DOI:** 10.1101/2025.04.15.25325855

**Authors:** Rachel E. Murray-Watson, Margherita Molaro, Rebecca J. Murray-Watson, Sakshi Mohan, Bingling She, Tara Mangal, Joseph H. Collins, Sangeeta Bhatia, Eva Janoušková, Timothy B. Hallett

## Abstract

Malawi is vulnerable to climate-related shocks, which are projected to worsen. Whilst some dimensions of this vulnerability have been characterised, little is known about healthcare sector resilience. Coupling facility-specific data on antenatal care (ANC) service provision in Malawi with gridded precipitation data, we use regression analyses to characterise the historic relationship between precipitation and healthcare access. We estimate that, between 2012 and 2024, precipitation negatively impacted ANC service utilisation in Malawi, with up to 1 in 20 appointments disrupted annually in some districts. Projecting further to 2060 indicates that, cumulatively, up to 250,000 pregnancies could be affected. Notably, if precipitation patterns from 1941–1953 had persisted into the 21st century, disruptions between 2012 — 2024 would be a hundred times less frequent, highlighting the significant influence of anthropogenic climate change on healthcare access. In a country already facing high maternal and neonatal mortality, such disruptions could further hinder access to care and worsen health outcomes.

## 2 Main

In the Intergovernmental Panel on Climate Change’s 2022 Report [1], Malawi was identified as one of the most vulnerable countries to climate change. Whilst this vulnerability has been explored in terms of food security [2, 3], economic activity [4, 5, 6], and some infectious diseases (e.g. malaria, [7]), little consideration has been given to the effect climate - and particularly, extreme weather events - has on the access to healthcare and health system functioning. Yet with many low-lying regions, unpaved roads, and poorly constructed buildings, Malawi’s health infrastructure is vulnerable to a variety of weather events, especially extreme precipitation, flooding, and landslides [7].

Indeed, between 1991 and 2020, more than 3.5 million people were exposed to flooding in Malawi, with 935 confirmed deaths related to flooding, e.g. due to mudslides, disease, and injury [7]. In addition to these direct impacts on individual health, precipitation events can severely disrupt access to and provision of medical care. In 2023, Cyclone Freddy affected patients’ access to 74 healthcare facilities, and hindered the operation of a further five [8], one of which was forced to close for 17 months [9]. The frequency of similar extreme weather events has been increasing in Malawi in recent decades, with more tropical cyclones recorded between 2012–2023 than occurred between 1946–2008 [7]. Given that Malawi’s average temperature has already increased 1.2^°^C over pre-industrial levels [10], such trends in extreme weather may be expected to persist as the climate continues to warm. Indeed, the World Bank posits that floods will be the most prominent climate-related threats Malawi faces [7]. However, since there is no systematic data collection on the impact of these and other weather-mediated disruptions on healthcare supply or access (such as road disruptions or a diminished healthcare workforce) the magnitude of their consequences for Malawi is unknown.

In other regions, some evidence linking healthcare access and adverse weather events has been found. Both the level of precipitation, the accessibility of a facility via paved roads - and the interaction between both - have been linked to reduced healthcare utilisation [11]. Flooding cuts off access to healthcare [12], and in regions such as Jakarta, Indonesia [13] and Assam, India [14], this exacerbates existing issues with healthcare supply. Fewer institutional deliveries occur in Mozambique’s rainy season compared to the dry season [15], whilst flooding events have been linked to lower utilisation of antenatal care (ANC) services in parts of Bangladesh [12].

There are limited such studies for Malawi: Cyclone Freddy was shown to reduce ANC utilisation in affected regions [16], though there is no estimation of the effect of cyclones or precipitation beyond this specific event, nor of the future impact. The majority of roads in Malawi are unpaved [17], and thus susceptible to heavy precipitation, providing a plausible mechanism of disruption. During Cyclone Freddy, roads in affected regions were rendered inaccessible due to flooding and landslides [18, 19], severely impinging on humanitarian responses. With the lack of transport to clinics already cited as a barrier to care among women seeking ANC services [20, 21], it is likely that unpaved roads’ vulnerability to precipitation - especially extreme precipitation - compounds the issue and drives down ANC utilisation even further [11, 15].

Here, we quantify the effects of precipitation on the utilisation of ANC services in Malawi. Compared to other healthcare services, ANC appointments are moderately time-constrained: definitionally they must precede the end of pregnancy. Therefore, disruptions to ANC services are comparatively more likely to cause missed appointments, rather than to delayed appointments (though, earlier in pregnancies, delays of a few weeks are possible). Additionally, given that future ANC service utilisation is primarily associated with demography, the demand for these services is simpler to project than that of, for example, infectious diseases. Importantly, attendance of ANC appointments is critical in reducing neonatal and maternal morbidity and mortality [22, 23, 24, 25], meaning that weather-mediated disruptions to these services could have severe health consequences. The utilisation of ANC services is a strong predictor of engagement with other maternity services, including institutional delivery and postnatal care. Consequently, disruptions in ANC services can serve as an indicator of broader healthcare service disruptions [26, 27].

Using facility-specific data on ANC service utilisation in Malawi [28] coupled with climate data from 2011 to 2024 [29], we use regression analyses to characterise the historic relationship between precipitation and ANC service delivery. We then use spatially downscaled climate model projections of precipitation for the period 2025-2060 [30] to estimate the impact across three divergent Shared Socioeconomic Pathways (SSPs). We estimate that, between 2012 and 2024, utilisation of ANC services was adversely affected by precipitation in Malawi, with up to 1 in 20 annual appointments disrupted in some districts. Future projections broadly replicate these historic patterns, potentially leading to a quarter of a million affected pregnancies between 2025 and 2060. Interestingly, if precipitation patterns from 1941-1953 had persisted into the early 21st century, there would be a hundred-fold fewer disruptions, indicating the consequential role of anthropogenic climate change in the provision of healthcare services. In a country with already-high maternal and neonatal mortality [31, 32, 33], such disruptions could increase barriers to care and worsen health outcomes.

## 3 Historical relationship between precipitation and ANC appointments

For our outcome metric, we use the total number of ANC services delivered per clinic each month. We obtained these data from Malawi’s District Health Information Software (DHIS2; [28]) and used records between January, 2011 and October, 2024 (see Section 7). For our precipitation variables, we used gridded reanalysis data for 2011–2025 [29], locating clinics to the appropriate grid square using a KDTree sorting algorithm [34]. From this we calculated several monthly precipitation metrics, informed by the Expert Team on Climate Change Detection and Indices (ETCCDI, [35]): the monthly maximum 5-day consecutive precipitation (Rx5day), the monthly cumulative precipitation (Figure 1 A), and various lags in these metrics.

**Figure 1:**
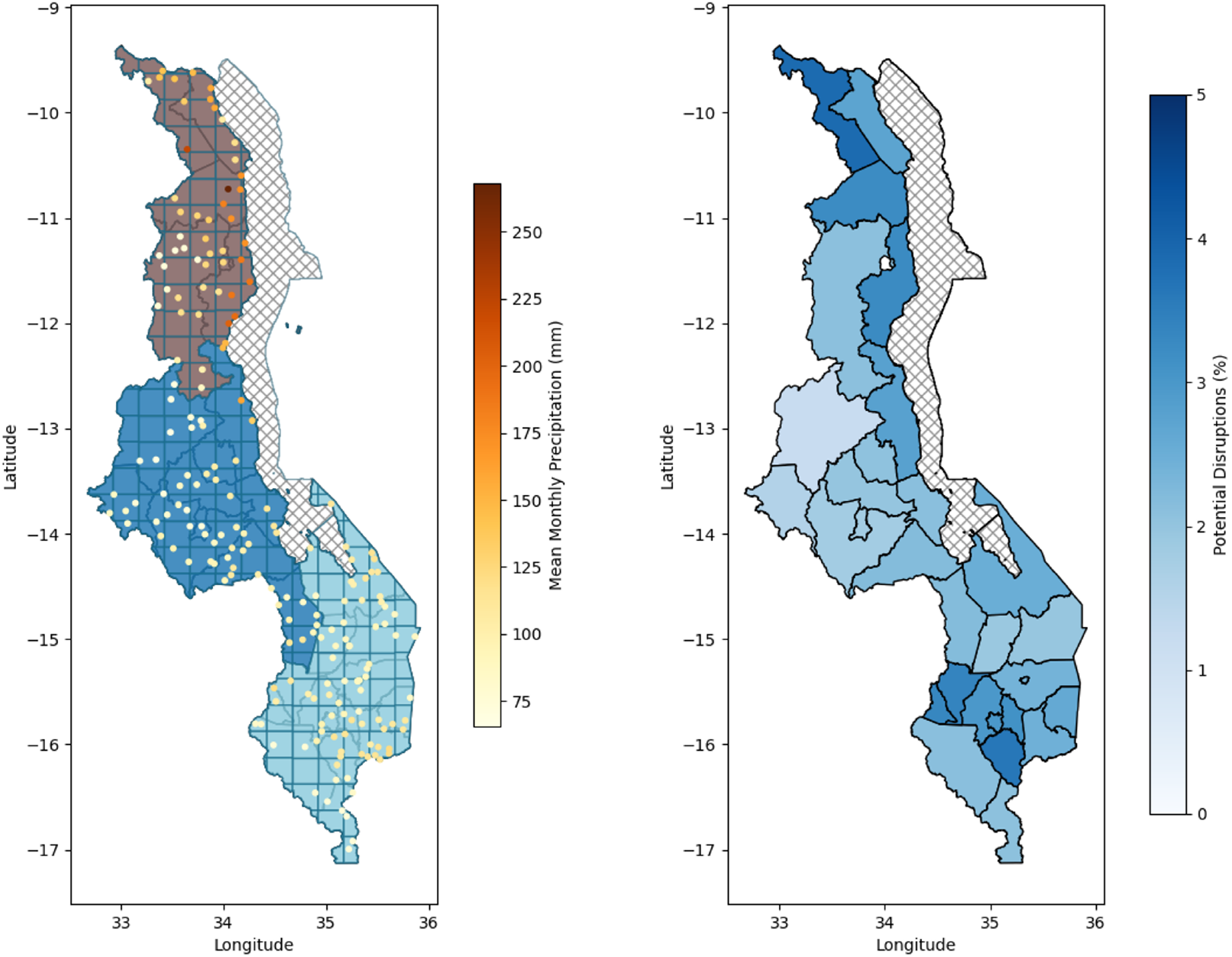
A. Mean monthly cumulative rainfall (mm) per month for 2011 to 2024. Each dot represents a facility, whilst its colour indicates the mean monthly precipitation for its grid square between 2011 and 2024. Each colour block represents a region (Northern, Central, and Southern Malawi). B. Percent of ANC visits disrupted by precipitation by district between 2011-2024.

We used backwards stepwise selection to create two linear regression models to predict future ANC visits (7.6, [15]): one without any precipitation indices (*M*_0_), and one including precipitation indices (*M*_1_). In *M*_0_, the setting of the facility (urban or rural), the owner of the facility (Government, Christian Health Associations of Malawi (CHAM), Non-Governmental Organisation (NGO), or Private), and its Euclidean distance to the next closest facility were significantly associated with counts of ANC appointments (Table 1, Section 7.6). For the precipitatin indices in *M*_1_, only the cumulative monthly precipitation was significant, though other metrics were included in the final, selected model.

**Table 1:**
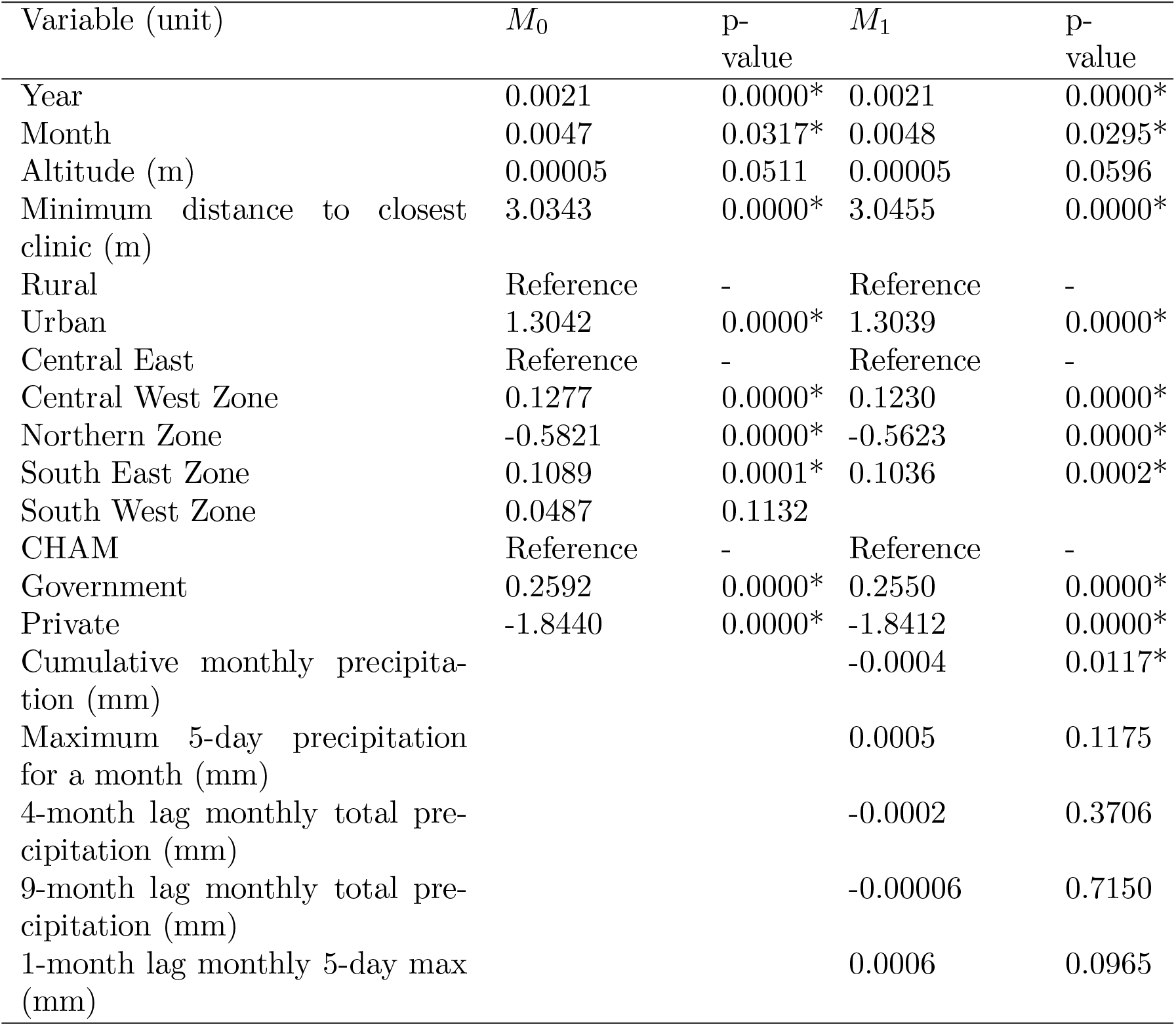
Regression results, including coefficients and p-values for various predictors, based on a models developed on data from 2011 to 2024. Starred p-values indicate significance at *p* < 0.05. Categorical data was sourced from Malawi’s Harmonized Health Facility Assessment [38], with precipitation indices calculated from the ERA5 reanalysis data [29].

The difference in predictions between these two models gave us our expected change in ANC services used due to precipitation (Figure 2, [15]). Though the coefficient for the cumulative monthly precipitation was small (−0.0004 ANC visits per facility per month per millimetre of monthly precipitation, Table 1), this translated into large effects with up to 4.7% of cases affected in some districts (mean across all districts of 2.5% 95% CI 2.25-2.73); Figure 1B). This translates to an estimated 66,000 affected pregnancies over 13 years, assuming that one live birth equates to around 4.4 ANC appointments delivered (accounting for stillbirths; [36, 37]).

**Figure 2:**
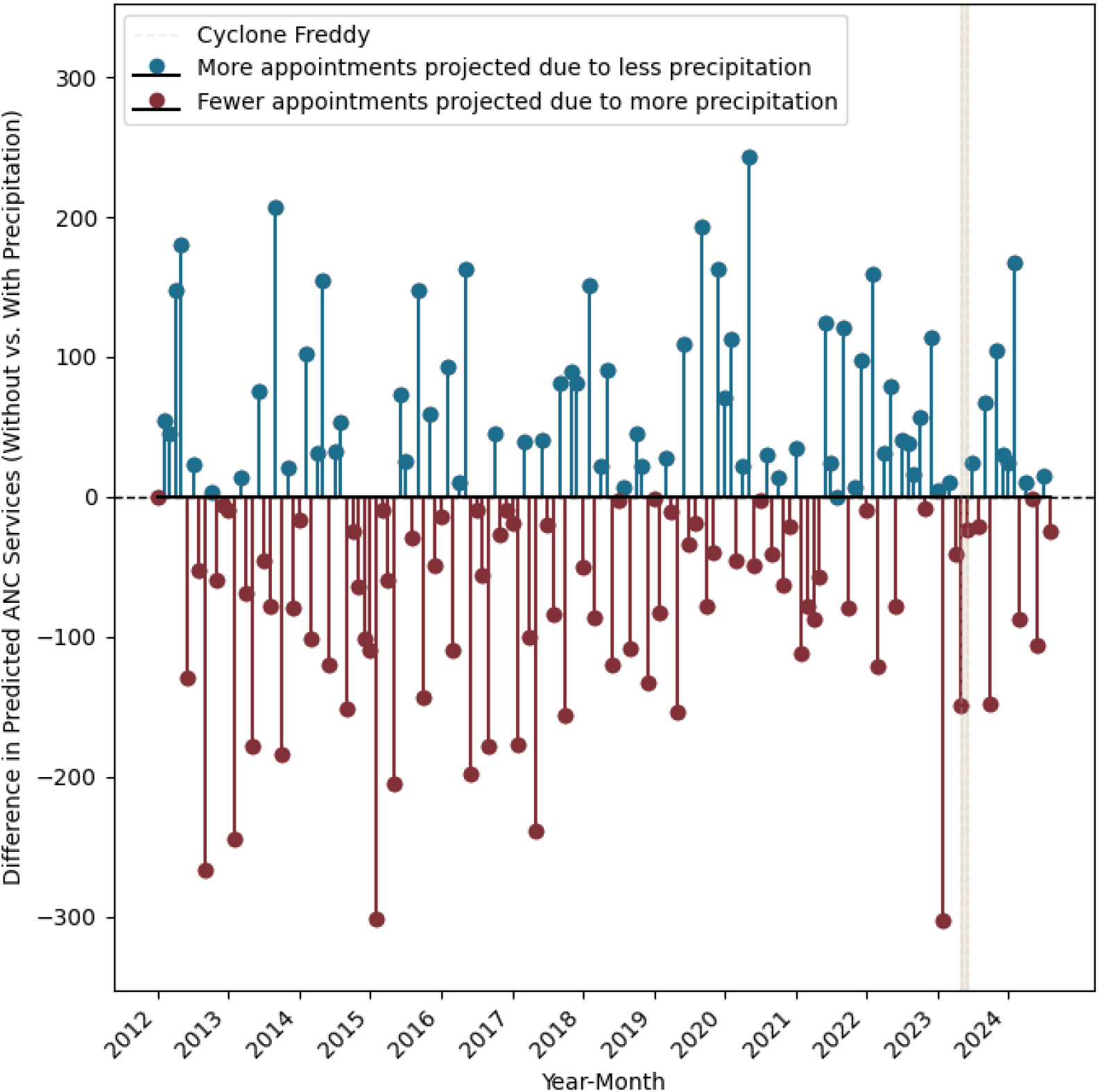
Monthly projected disruption of ANC service delivery across Malawi, calculated as the difference between models *M*_0_ and *M*_1_ (without and with precipitation indices respectively) for years 2011–2024. The beige bar indicates months affected by Cyclone Freddy (February and March, 2023).

## 4 Projecting forward: future disruptions under different climate scenarios

We considered three divergent future scenarios, based on the Shared Socioeconomic Pathways (SSP) framework [39] (Section 7): SSP1-2.6, SSP2-4.5, SSP5-8.5. These correspond to warming temperatures, globally, of approximately 1.7 C, 2 ^°^C, and 2.4 ^°^C, respectively, by 2060, our horizon of interest. Using projections on future pregnancies from the Thanzi La Onse (tlomodel.org) model, a Malawi-specific, individual-based, multi-disease and healthcare system model, (Section see7) [40, 37], we found that up to a quarter of a million pregnancies could be impacted by precipitation by 2060.

Irrespective of the scenario and model used, districts directly bordering Lake Malawi and in the south suffered the most disruption (Figure 3), areas which have been highlighted as particularly vulnerable to future flooding and landslides [7].

**Figure 3:**
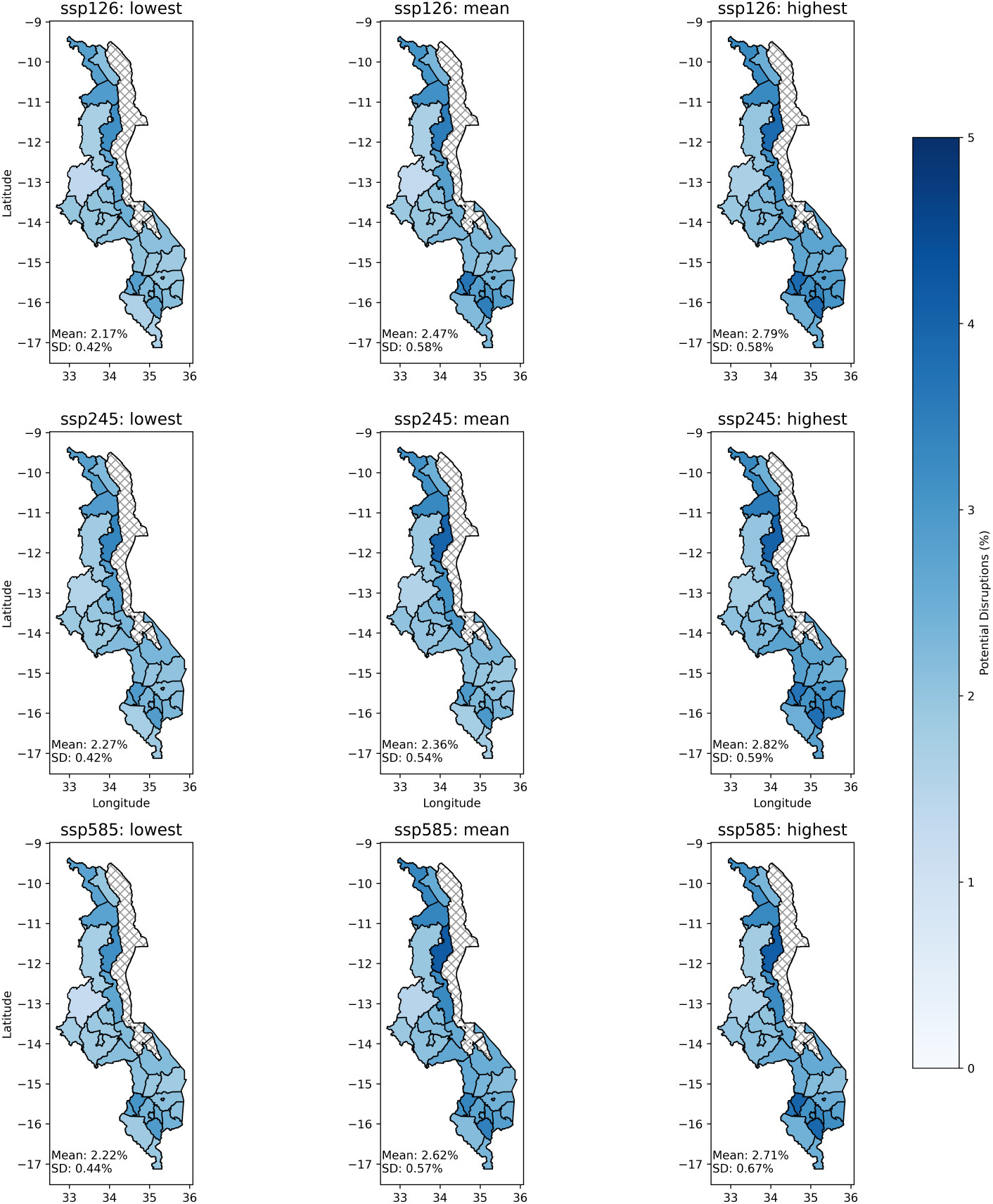
Projected percentage of ANC services potentially disrupted by precipitation events between 2025 and 2060.

The cumulative magnitude of disruptions is similar across all scenarios (Table 2). This may be due to the effects of disparate effects during the wet and dry seasons [15, 10]. Though extreme precipitation events are likely to increase, so too are periods of little to no rainfall [7]. Given that our model indicates that increase in monthly precipitation is associated with a decrease in ANC services, extended dry spells could mean periods of little transport or road disruptions. That is not to say that drought will have positive, or even neutral, effects on ANC utilisation, but they would need to be accounted for explicitly in a model.

**Table 2:**
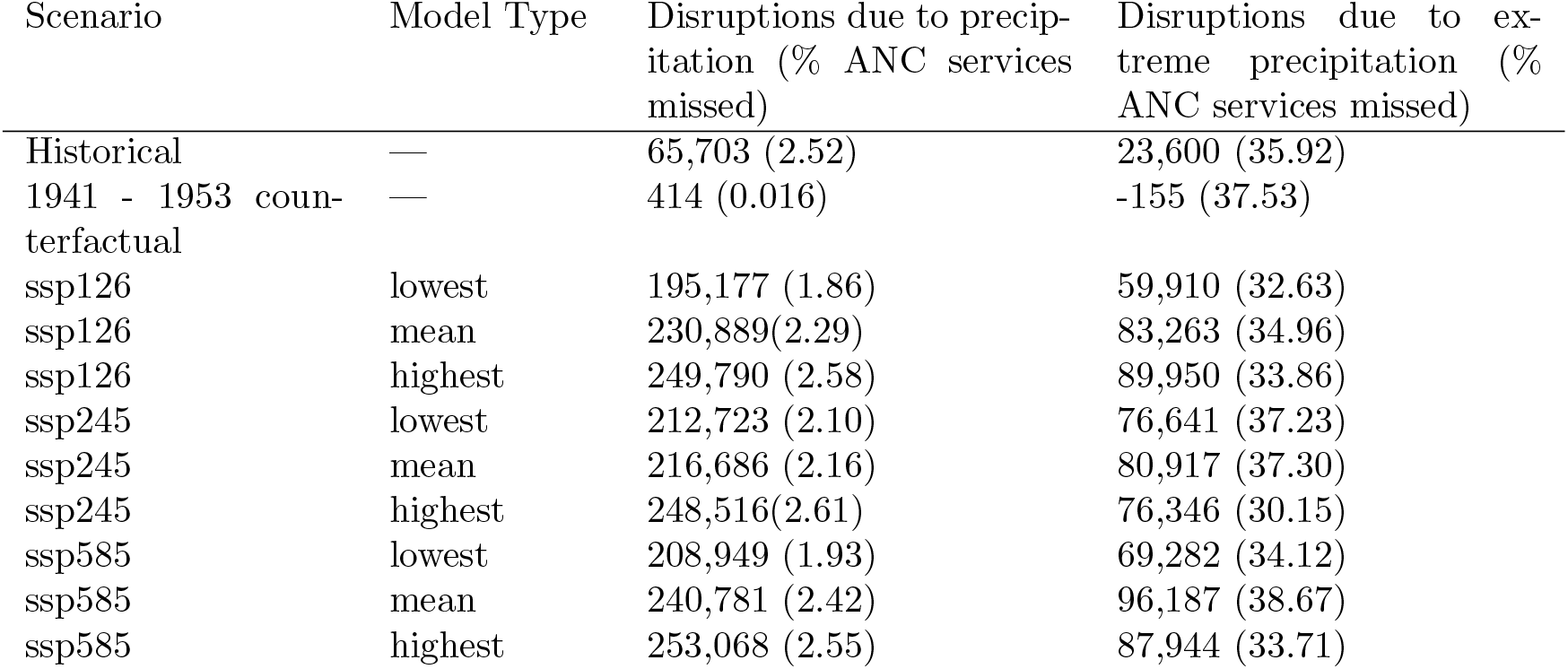
Cumulative disruption in ANC visits between in the “historical” (2012-2024), counterfactual, and 2025-2060 scenarios, nationally. Extreme precipitation is based on the 90th percentile of the historical rainfall between 2011-2024 (318 mm per month).

| Scenario | Model Type | Disruptions due to precipitation (% ANC services missed) | Disruptions due to extreme precipitation (% ANC services missed) |
| --- | --- | --- | --- |
| Historical | — | 65,703 (2.52) | 23,600 (35.92) |
| 1941 - 1953 counterfactual | — | 414 (0.016) | -155 (37.53) |
| ssp126 | lowest | 195,177 (1.86) | 59,910 (32.63) |
| ssp126 | mean | 230,889 (2.29) | 83,263 (34.96) |
| ssp126 | highest | 249,790 (2.58) | 89,950 (33.86) |
| ssp245 | lowest | 212,723 (2.10) | 76,641 (37.23) |
| ssp245 | mean | 216,686 (2.16) | 80,917 (37.30) |
| ssp245 | highest | 248,516 (2.61) | 76,346 (30.15) |
| ssp585 | lowest | 208,949 (1.93) | 69,282 (34.12) |
| ssp585 | mean | 240,781 (2.42) | 96,187 (38.67) |
| ssp585 | highest | 253,068 (2.55) | 87,944 (33.71) |

What is notable, however, is that in the more extreme climate scenarios, a higher percentage of disruptions are due to extreme precipitation events (318 mm over one month, which is the 90th percentile of historical rainfall between 2011-2024; Table 2) compared to the “moderate” SPP1-2.6. Even when the overall proportion of pregnancies affected by precipitation remains constant between scenarios, extreme events cause the most disruption in SSP5-8.5.

## 5 A different baseline: precipitation trends from 1940-1950

It is evident from our analysis that the patterns of ANC disruption between 2010–2024 are largely carried forward to 2060, irrespective of variations SSP. This may be attributable to the existing effects of climate change, which have already induced profound changes in Malawi’s climate [7]. Therefore, to understand the potential effect of anthropogenic climate change on ANC disruptions, we must use precipitation data from an earlier period.

We used ERA5 reanalysis precipitation data for 1941-1953 as a counterfactual data for our *M*_1_ model for 2011-2024. Under this scenario, we find a much reduced impact of precipitation on the disruption of ANC visits: only 414 (0.016%) of pregnancies suffered any disruption, which is 157-fold fewer than the “factual” (“historical”) scenario. The pattern of geographical disruption was preserved.

## 6 Discussion

Healthcare resilience under a changing climate is a topic of emerging concern [41, 10]. To date, little work has been done to understand what risks extreme weather events pose to healthcare access, particularly in low- and middle-income countries. Here, we show that antenatal care (ANC) service delivery in Malawi is adversely affected by precipitation events, particularly extreme events.

We estimate that, between 2012 and 2024, around 2.5% of pregnancies in Malawi were affected by precipitation-mediated disruptions to ANC care (assuming that for each live birth, there have been 4.4 ANC appointments per pregnant women). In the coming decades up to 2060, between 1.9 and 2.6% of ANC services will be disrupted, depending on the SSP scenario, translating into approximately 195,000 and 253,000 affected pregnancies. The cumulative projected disruptions, therefore, largely match historical patterns, potentially due to the impact of existing climate change [39], the impacts of which are already being felt, and will continue to be felt for decades. However, using precipitation data from 1941-1953, there is a marked reduction in the number of disruptions, with fewer than 0.02% of the pregnancies affected, suggesting what climate change has occurred between 1953 and 2024 has negatively impacted ANC service utilisation in Malawi.

Across all scenarios, the broad similarity in national trends between scenarios belies regional and temporal variation. Whilst the mean cumulative potential disruption by district was 2.5% (standard deviation of 0.64%), districts that are in the South or bordering Lake Malawi may experience disruptions of up to 1 in 20 annual appointments, approximately 33% worse than historical patterns.

An additional potential reason for the lack of inter-scenario variability is because, though higher temperatures increase the likelihood of extreme precipitation events, they do so too for drought. We have treated precipitation as a continuous variable, with no minimum threshold for disruption, thus allowing for the possibility that whilst there will be more extreme events that could cause major disruptions, if “normal” precipitation events will become less common, these lower-level disruptions will disappear. Though extended periods of low precipitation, or even drought, are implicitly included in our analyses at a monthly resolution, the further inclusion of drought metrics as defined by the ETCCDI would provide further insight into this relationship.

In other “impact attribution” studies, where the consequences of extreme weather events are probabilistically associated with climate change [42], the typical baseline “pre-anthropogenic climate change” climate is taken as 1850-1900, before the onset of rapid industrialisation and use of fossil fuels. However, in our ERA5 reanalysis dataset, the earliest available data are from 1940. Despite this limitation, data from this period still precede the observed rise in extreme precipitation events [10], making it a reasonable baseline for our analysis.

Previous work in this area is limited, but estimates of ANC provision across Malawi in the six weeks preceding and succeeding Cyclone Freddy in a subset of clinics in the South West of Malawi showed a 60% reduction in ANC usage [16]. Though our analysis works at a different resolution, and predicts based on historical trends rather than the immediate pre- and post-event periods, we note that for February 2023 there was an estimated mean disruption to ANC services of approximately 5% across the entire region, with facility-level disruptions of up to 19%.

It is possible that we are under-estimating the level of disruptions. The monthly resolution of our data means that if an appointment is delayed, but occurs later in the same calendar month, it is not recorded as disrupted. Additionally, though we do account for the Euclidean distance to the closest clinic in our models, women may travel to further clinics, or preferentially travel to those that are accessible by paved roads or other transport links. These compensatory mechanisms were not possible to discern with the data we had available to us. Conversely, by not accounting for potential improvements in infrastructure, we could over-estimate the negative impact of precipitation on healthcare provision and utilisation.

This work only considers the simple relationship between precipitation and utilisation of ANC services. Whilst it is possible that a large part of the effect of precipitation is mediated by reduced accessibility of healthcare facilities, we have not included it explicitly via models of flooding and/or road networks, which - based on other regional studies - are likely to be mediators of the relationship that we have quantified.

Data scarcity meant that only a subset of all healthcare facilities in Malawi were included in our study (26.2% of all facilities expected to report on ANC services by DHIS2), meaning that smaller and potentially more vulnerable sites have been omitted. Furthermore, in resource-limited environments, if an extreme weather event, routine ANC services may be de-prioritised in favour of more urgent needs. Neither of these effects are accounted for in our model.

It is also possible that the disruptions we observe are because precipitation and associated flooding will limit the provision of healthcare. Just as patients may have difficulty accessing clinics, so too may healthcare providers, and supply chains disruptions may result in consumable stock-outs. As happened in Cyclone Freddy, healthcare clinics may be damaged and/or destroyed, limiting or eliminating the provision of care.

Even with these limitations, our analysis demonstrates that the upward trend in precipitation levels, particularly when linked to extreme events, will have negative consequences for the utilisation of ANC services in Malawi. Given the link between access to these services and improved maternal and neonatal outcomes [25], such disruptions could have severe consequences for health if left unaddressed.

## 7 Methods

### 7.1 ANC Data

We obtained data on the monthly number of antenatal care (AntenatalTotal) visits from Malawi’s District Health Information Systems (DHIS2) [28]. These data are available monthly, setting the temporal resolution of our analysis. After cross-referencing with clinics for which we had data available from Malawi’s HMalawi’s Harmonized Health Facility Assessment [38], and omitting clinics where ¿90% of observations were missing, we obtained ANC visit data for 199 healthcare facilities (26.2 % of the rural and urban hospitals, healthcare clinics, and health centres expected to report data to DHIS2) from 2011 and 2024. Excluding missing data in the remaining clinics, we have 21,133 observations for this time period. For details on how we dealt with missing data, see Appendix 7.5.

### 7.2 Climate Data

We obtained daily historical precipitation levels in Malawi from ERA5, a gridded reanalysis product produced by European Centre for Medium-Range Weather Forecasts (ECMWF) [29]. We obtained data from 2011-2024, and used it to calculate the cumulative monthly precipitation for each month in that period and 1,2,3,4, and 9-month lag in precipitation. Additionally, we calculated the 5-day maximum cumulative precipitation for each month [35], and included the same lag periods as the monthly data. The square of the cumulative monthly and 5-day max indices were included, alongside the interaction between them.

For estimates of future precipitation, we used downscaled Coupled Model Inter-comparison Project phase 6 from the Carbon Impact Lab [30]. These data are at a 1/4-degree regular global grid and use ERA5 as a reference dataset. We examined three Shared Socio-Economic Pathways (SSPs), which represent different scenarios for the future climate depending on societal, demographic, and economic trends [39]. These scenarios were SSP1-1.9, which would result from sustainable growth and increased societal equality; SSP2-4.5, the “middle of the road” which broadly follows historical patterns across all factors of interest, and SSP5-5.8, which results from increased fossil-fuel driven development.

To avoid averaging out extremes in precipitation events, for each of the scenarios, we chose the model projections with the lowest, mean, and highest mean monthly prediction over all grid squares in the 35-year period of interest (Table 3). We used the KDTree algorithm to assign each clinic to the appropriate grid square for analysis.

**Table 3:** Downscaled CMIP6 models comprising the model ensemble in our analysis.

| Scenario | Lowest | Median | Highest |
| --- | --- | --- | --- |
| SSP1-1.9 | HadGEM3-GC31-LL | MPI-ESM1-2-LR | INM-CM5-0 |
| SSP2-4.5 | HadGEM3-GC31-LL | UKESM1-0-LL | INM-CM5-0 |
| SSP5-8.5 | HadGEM3-GC31-LL | EC-Earth3-CC | CMCC-CM2-SR5 |

### 7.3 Covariate Data

In our initial model fitting, we included the following covariates: the precipitation indices (cumulative monthly rainfall (mm), 5-day maximum monthly rain-fall (mm), the 1-,2-,3-,4-,9-month lag in these metrics (mm), and their squares (mm^2^), cubes (mm^3^) and products (mm^2^), the year and month of observation, the facility ID, its zone (Northern, Central East, Central West, South East, South West), type (Dispensary, Health Centre, Clinic, Rural/Community Hospital, District Hospital), district, and owner (Government, Christian Health Associations of Malawi, NGO, or Private), whether it was urban or rural, the distance to the closest facility (m), and its altitude (m).

A covariance matrix indicated a high level of correlation between the facility owner and the facility type; we therefore removed the latter from the predictions. Similarly, zone was highly correlated with the district, so we removed the latter.

### 7.4 Projections of pregnancy

We used the TLO model (tlomodel.org, [43]) to estimate the number of pregnancies for the years 2011-2060. TLO is an individual-based, all-disease, whole health-system model specific to Malawi, though at present, these projections are made in the absence of information on climate change. Therefore, the birth projections are constant across all future scenarios. Additionally, in order to generate projections on births, we use an idealised scenario in which there are no limitations on the availability of or access to healthcare (i.e., there are no limitations on the availability of consumables and equipment, healthcare workers can operate to their fullest capacity, and those who need medical attention seek it) [40, 37].

### 7.5 Missing Data from the Electronic Health Records

There appeared to be a pattern of non-randomness in the DHIS2 entries around the time of Cyclone Freddy in Malawi (February-March 2023). In particular, when news reports [9] confirmed that a Phalombe Clinic was closed due to cyclone damage, entries in DHIS2 were missing, rather than “0” (indicating that no services were delivered). Yet for Nkhulabme Health Centre, which was also closed due to damage, “0” services were returned to DHIS2. A third clinic, which is in close proximity with these two clinics - the Thumbwe Health Centre - also reported missing data, suggesting it too was affected by the cyclone, though no news reports could confirm this.

To account for this, where secondary sources could confirm the closure or otherwise of a clinic, the missing data were replaced with a “0” (indicating no services were delivered). However, where such sources could not be confirmed, we assumed that the lack of data was due to random non-reporting, and not associated with weather.

Missing data on altitude were imputed based on the mean altitude of all clinics.

Likoma had no clinics with the requisite covariate data, so was omitted from the analyses.

### 7.6 Analysis

We constructed two models to predict the future number of antenatal care (ANC) services delivered each month in each clinic: one with no information on precipitation, and one with. Both of these were linear regression models, using logged counts of ANC services for each month as our outcome variable.

We used backward stepwise selection to choose the best model based on Akaike information criterion (AIC), resulting in the following:

Model *M*_0_, without precipitation indices (AIC = 63501):

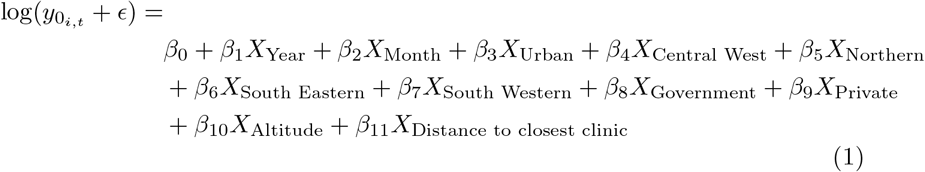

where *ϵ* = 1 to allow for the calculation of logs when the number of ANC services delivered are 0.

The model *M*_1_ with precipitation indices is as follows (Δ*AIC* = −6):

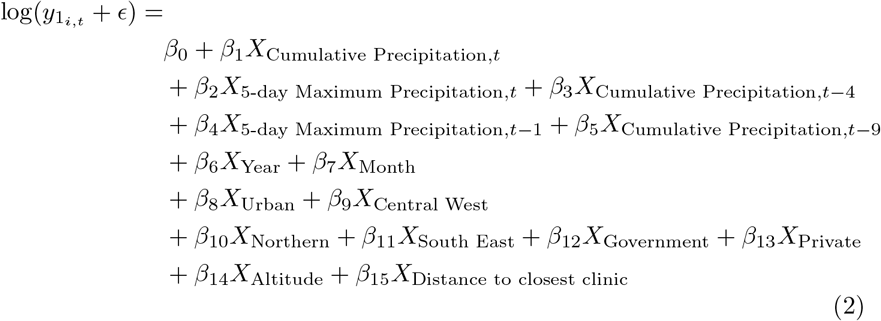

In both models, categorical variables (facility ownership (Government, Non-Governmental Organisation (NGO), or Private), Region (Northern, Central Western, Central Eastern, South Eastern, South Western) and location (Urban or Rural)) were one-hot encoded to control for their effects.

Coefficients and p-values for these models are shown in Tables 1 – **??**. Both models had a high adjusted *R*^2^ (> 0.95).

We considered the difference in the predictions of these two models to be the number of ANC services that were disrupted due to precipitation for month *i* in year *j* (Equation 3, where the 0 subscript indicates no precipitation indices, and 1 their inclusion) [15]. We reason that the omission of precipitation indices will give us the expected number of ANC services delivered assuming that precipitation has no effect. Therefore, if inclusion of the indices changes the predicted ANC services delivered, we attribute that difference to precipitation. A likelihood ratio test indicated that model *M*_1_ was better at predicting the data than the *M*_0_, precipitation-free model (LR: 14.0 on 4 degrees of freedom, p-value = 0.007).

As we are primarily interested in the negative disruptions of precipitation events, in presenting our results we will only consider months where the “precipitation” model predicts a deficit of ANC services delivered. Given the monthly resolution of our data, we were not able to capture any appointments that were postponed by a few days but still fell within the recording month of interest: only ones that were missed entirely, or shifted to the next month (inflating those figures) could be captured. Our inclusion of lagged precipitation metrics helps to account for the latter effect.

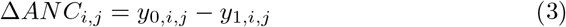

## Data Availability

All data produced in the present study are available upon reasonable request to the authors
The climate data is available from ECMWF.

https://cds.climate.copernicus.eu/datasets/reanalysis-era5-pressure-levels-monthly-means?tab=overview

## References

[1] J. Birkmann, E. Liwenga, R. Pandey, E. Boyd, R. Djalante, F. Gemenne, W. Leal Filho, P. F. Pinho, L. Stringer, and D. Wrathall. Poverty, livelihoods and sustainable development. In H.-O. Pörtner, D. C. Roberts, M. Tignor, E. S. Poloczanska, K. Mintenbeck, A. Alegría, M. Craig, S. Langsdorf, S. Löschke, V. Möller, A. Okem, and B. Rama, editors, Climate Change 2022: Impacts, Adaptation and Vulnerability, pages 1171– 1274. Cambridge University Press, Cambridge, UK and New York, NY, USA, 2022.

[2] Erika A Warnatzsch and David S Reay. Assessing climate change projections and impacts on central malawi’s maize yield: The risk of maladaptation. Science of the Total Environment, 711:134845, 2020.

[3] Muhammad Abid, Akhter Ali, Mohsin Raza, Mubashir Mehdi, et al. Ex-ante and ex-post coping strategies for climatic shocks and adaptation determinants in rural malawi. Climate Risk Management, 27:100200, 2020.

[4] Karl Pauw, James Thurlow, and Dirk Van Seventer. Droughts and floods in malawi assessing the economywide effects, 2010, 2022.

[5] Channing Arndt, Paul Chinowsky, Charles Fant, Sergey Paltsev, C Adam Schlosser, Kenneth Strzepek, Finn Tarp, and James Thurlow. Climate change and developing country growth: the cases of malawi, mozambique, and zambia. Climatic Change, 154:335–349, 2019.

[6] Askar Mukashov, Timothy Thomas, and James Thurlow. Revisiting development strategy under climate uncertainty: case study of malawi. Climatic Change, 177(6):1–18, 2024.

[7] World Bank. Malawi - climate and health vulnerability assessment. http://hdl.handle.net/10986/41847, 2024. xLicense: CC BY-NC 3.0 IGO.

[8] Prosper Lutala and Martha Makwero. Cyclone freddy in malawi: Reflections from a primary care perspective. African Journal of Primary Health Care & Family Medicine, 15(1):1–2, 2023.

[9] Malawi News Agency. Phalombe health centre re-opened for operation, 2025. Accessed: 2025-01-14.

[10] World Bank. Climate change knowledge portal, 2024. Accessed: 2024-10-01.

[11] Prestige Tatenda Makanga, Nadine Schuurman, Charfudin Sacoor, Helena Edith Boene, Faustino Vilanculo, Marianne Vidler, Laura Magee, Peter von Dadelszen, Esperanca Sevene, Khatia Munguambe, et al. Seasonal variation in geographical access to maternal health services in regions of southern mozambique. International journal of health geographics, 16:1–16, 2017.

[12] Hilde Orderud, Juho Härkönen, Cathrine Tranberg Hårsaker, and Malin Bogren. Floods and maternal healthcare utilisation in bangladesh. Population and environment, 44(3):193–225, 2022.

[13] Isabell G Klipper, Alexander Zipf, and Sven Lautenbach. Flood impact assessment on road network and healthcare access at the example of jakarta, indonesia. AGILE: GIScience Series, 2:4, 2021.

[14] Jayanti Saha. The pattern of morbidity and access to healthcare service in the riverine flood-prone villages of assam, india. The Open Public Health Journal, 16(1), 2023.

[15] Briana Stone, Julia Sambo, Talata Sawadogo-Lewis, and Timothy Roberton. When it rains, it pours: detecting seasonal patterns in utilization of maternal healthcare in mozambique using routine data. BMC health services research, 20:1–10, 2020.

[16] Hussein H Twabi, James Jafali, Leonard Mndala, Jennifer Riches, Edward JM Monk, Deborah Phiri, Regina Makuluni, Luis Gadama, Fannie Kachale, Rosemary Bilesi, et al. Cyclone freddy and its impact on maternal health service utilisation: Cross-sectional analysis of data from a national maternal surveillance platform in malawi. PLOS Global Public Health, 4(8):e0003565, 2024.

[17] Roads Authority Malawi. Road network in malawi, n.d. Accessed: 2025-01-20.

[18] ReliefWeb. Situation report. malawi tropical cyclone freddy: Transport and logistics cluster situation, 2023. Available: here [Accessed 20 Jan 2025].

[19] Malawi Meteorological Department. State of malawi climate 2023, 2023. Accessed: 2025-01-20.

[20] L Manda-Taylor, DA Sealy, and Joni Roberts. Factors associated with delayed antenatal care attendance in malawi: results from a qualitative study. Medical Journal of Zambia, 44(1):17–25, 2017.

[21] Chancy S Chimatiro, Precious Hajison, Effie Chipeta, and Adamson S Muula. Understanding barriers preventing pregnant women from starting antenatal clinic in the first trimester of pregnancy in ntcheu district-malawi. Reproductive health, 15:1–7, 2018.

[22] World Health Organization et al. Who recommendations on antenatal care for a positive pregnancy experience. In WHO recommendations on antenatal care for a positive pregnancy experience, pages 152–152. 2016.

[23] Amsalu Taye Wondemagegn, Animut Alebel, Cheru Tesema, and Worku Abie. The effect of antenatal care follow-up on neonatal health outcomes: a systematic review and meta-analysis. Public health reviews, 39:1–11, 2018.

[24] Tesfalidet Tekelab, Catherine Chojenta, Roger Smith, and Deborah Loxton. The impact of antenatal care on neonatal mortality in sub-saharan africa: A systematic review and meta-analysis. PloS one, 14(9):e0222566, 2019.

[25] Getahun Tiruye, Kasiye Shiferaw, Addisu Shunu, Yitagesu Sintayeu, and Abdulbasit Musa Seid. Antenatal care predicts neonatal mortality in eastern africa: a systematic review and meta-analysis of observational studies. Journal of Neonatology, 36(1):42–54, 2022.

[26] Gedefaw Abeje Fekadu, Getachew Mullu Kassa, Abadi Kidanemariam Berhe, Achenef Asmamaw Muche, and Nuradin Abusha Katiso. The effect of antenatal care on use of institutional delivery service and postnatal care in ethiopia: a systematic review and meta-analysis. BMC health services research, 18:1–11, 2018.

[27] Alehegn Bishaw Geremew, Moges Muluneh Boke, and Ayenew Engida Yismaw. The effect of antenatal care service utilization on postnatal care service utilization: A systematic review and meta-analysis study. Journal of pregnancy, 2020(1):7363242, 2020.

[28] Malawi HMIS. Malawi health management information system. https://dhis2.health.gov.mw/, 2022. xAccessed: 2023-10-25.

[29] Hans Hersbach, Bill Bell, Paul Berrisford, Shoji Hirahara, András Horányi, Joaquín Muñoz-Sabater, Julien Nicolas, Carole Peubey, Raluca Radu, Dinand Schepers, et al. The era5 global reanalysis. Quarterly Journal of the Royal Meteorological Society, 146(730):1999–2049, 2020.

[30] Diana Gergel, Kelly McCusker, Brewster Malevich, Emile Tenezakis, Meredith Fish, and Michael Delgado. Climateimpactlab/downscalecmip6: (v1.0.0). 10.5281/zenodo.6403794, 2022. Zenodo.

[31] Leontine Alkema, Doris Chou, Daniel Hogan, Sanqian Zhang, Ann-Beth Moller, Alison Gemmill, Doris Ma Fat, Ties Boerma, Marleen Temmerman, Colin Mathers, et al. Global, regional, and national levels and trends in maternal mortality between 1990 and 2015, with scenario-based projections to 2030: a systematic analysis by the un maternal mortality estimation inter-agency group. The lancet, 387(10017):462–474, 2016.

[32] World Health Organization. Trends in Maternal Mortality 2000 to 2017: Estimates by WHO, UNICEF, UNFPA, World Bank Group and the United Nations Population Division. World Health Organization, Geneva, 2019. Licence: CC BY-NC-SA 3.0 IGO.

[33] Katherine R Paulson, Aruna M Kamath, Tahiya Alam, Kelly Bienhoff, Gdiom Gebreheat Abady, Jaffar Abbas, Mohsen Abbasi-Kangevari, Hedayat Abbastabar, Foad Abd-Allah, Sherief M Abd-Elsalam, et al. Global, regional, and national progress towards sustainable development goal 3.2 for neonatal and child health: all-cause and cause-specific mortality findings from the global burden of disease study 2019. The Lancet, 398(10303):870– 905, 2021.

[34] Jon Louis Bentley. Multidimensional binary search trees used for associative searching. Communications of the ACM, 18(9):509–517, 1975.

[35] CLIVAR. Etccdi climate indices data, 2025. Accessed: 2025-01-13.

[36] National Statistical Office NSO/Malawi and ICF. Malawi Demographic and Health Survey 2015-16. NSO and ICF, Zomba, Malawi, 2017.

[37] Joseph Collins, Helen Allott, Wingston Ng’ambi, Ines Li Lin, Mosé Giordano Matthew Graham, Eva Janoušková, Fannie Kachale, Kondwani Kawaza, Tara Mangal, et al. Estimating the impact of maternity service delivery on health in malawi: An individual-based modelling study. 2024.

[38] Malawi - harmonized health facility assessment: 2018-2019 report (vol. 1 of 4): Main report. Technical Report 156080, World Bank Group, Washington, D.C., January 2019. Disclosed on January 24, 2021.

[39] Hoesung Lee, Katherine Calvin, Dipak Dasgupta, Gerhard Krinner, Aditi Mukherji, Peter Thorne, Christopher Trisos, Josèe Romero, Paulina Aldunce, Ko Barret, et al. Ipcc, 2023: Climate change 2023: Synthesis report, summary for policymakers. contribution of working groups i, ii and iii to the sixth assessment report of the intergovernmental panel on climate change [core writing team, h. lee and j. romero (eds.)]. ipcc, geneva, switzerland. 2023.

[40] Tim Colbourn, Eva Janoušková, Ines Li Lin, Joseph Collins, Emilia Connolly, Matt Graham, Britta Jewel, Fannie Kachale, Tara Mangal, Gerald Manthalu, et al. Modeling contraception and pregnancy in malawi: a thanzi la onse mathematical modeling study. Studies in family planning, 54(4):585–607, 2023.

[41] World Health Organization. WHO guidance for climate-resilient and environmentally sustainable health care facilities. World Health Organization, 2020.

[42] Donna James et al. Detection & attribution of climate change impacts on human health. Wellcome Open Res, 9(245):245, 2024.

[43] Timothy B Hallett, Tara D Mangal, Asif U Tamuri, Nimalan Arinaminpathy, Valentina Cambiano, Martin Chalkley, Joseph H Collins, Jonathan Cooper, Matthew S Gillman, Mose Giordano, et al. Estimates of resource use in the public-sector health-care system and the effect of strengthening health-care services in malawi during 2015–19: a modelling study (thanzi la onse). The Lancet Global Health, 13(1):e28–e37, 2025.

